# Mapping each pre-existing condition’s association to short-term and long-term COVID-19 complications

**DOI:** 10.1101/2020.12.02.20242925

**Authors:** AJ Venkatakrishnan, Colin Pawlowski, David Zemmour, Travis Hughes, Akash Anand, Gabriela Berner, Nikhil Kayal, Arjun Puranik, Ian Conrad, Sairam Bade, Rakesh Barve, Purushottam Sinha, John C. O’Horo, Andrew D. Badley, Venky Soundararajan

## Abstract

Understanding the relationships between pre-existing conditions and complications of COVID-19 infection is critical to identifying which patients will develop severe disease. Here, we leverage 1.1 million clinical notes from 1,903 hospitalized COVID-19 patients and deep neural network models to characterize associations between 21 pre-existing conditions and the development of 20 complications (e.g. respiratory, cardiovascular, renal, and hematologic) of COVID-19 infection throughout the course of infection (i.e. 0-30 days, 31-60 days, and 61-90 days). Pleural effusion was the most frequent complication of early COVID-19 infection (23% of 383 complications) followed by cardiac arrhythmia (12% of 383 complications). Notably, hypertension was the most significant risk factor associated with 10 different complications including acute respiratory distress syndrome, cardiac arrhythmia and anemia. Furthermore, novel associations between cancer (risk ratio: 3, p=0.02) or immunosuppression (risk ratio: 4.3, p=0.04) with early-onset heart failure have also been identified. Onset of new complications after 30 days is rare and most commonly involves pleural effusion (31-60 days: 24% of 45 patients, 61-90 days: 25% of 36 patients). Overall, the associations between pre-COVID conditions and COVID-associated complications presented here may form the basis for the development of risk assessment scores to guide clinical care pathways.

## Introduction

The COVID pandemic remains an ongoing public health crisis^1^, and it is critically important to understand the full spectrum of complications that arise throughout the course of COVID infection. There are already several emerging reports of risk factors of severe disease as well as lingering long-term effects such as fatigue, myalgia and kidney related complications^2^. However, there is an incomplete understanding of the relationship between pre-existing comorbidities and post-COVID complications.

Longitudinal multi-center patient data in EHRs of over 20,000 COVID-19 patients (1,903 hospitalized) from the Mayo Clinic (Rochester, Arizona, Florida) and associated health systems, provides a unique opportunity to understand the relationship between comorbidities and COVID complications^3^. While the structured EHR fields such as ICD codes are modestly informative, the true context of comorbidities and complications are buried in the 1.1 million+ unstructured patient notes of the 1,903 COVID-19 patients. In this study, we have leveraged ‘augmented curation’ of EHR notes in COVID patients^3^ to map the relationships between complications, comorbidities, and outcomes in the hospitalized COVID-19 patients.

## Methods

### Institutional Review Board (IRB)

This retrospective research was conducted under IRB 20–003278, ‘Study of COVID-19 patient characteristics with augmented curation of Electronic Health Records (EHR) to inform strategic and operational decisions’. For further information regarding the Mayo Clinic Institutional Review Board (IRB) policy, and its institutional commitment, membership requirements, review of research, informed consent, recruitment, vulnerable population protection, biologics, and confidentiality policy, please refer to www.mayo.edu/research/institutional-review-board/overview.

### Study design

In this study, we consider all hospitalized COVID-19 positive patients (positive PCR for SARS-CoV2) in the Mayo Clinic electronic health record (EHR) database from March 12, 2020 to September 15, 2020 (1,903 patients total). An overview of the clinical characteristics of this study population is provided in **Table 1**.

**Table 1:** General clinical characteristics of study population. Clinical characteristics of all hospitalized COVID-19 positive patients in the Mayo Clinic EHR dataset. For each clinical covariate, the number of unique patients in the dataset is shown, with the percentage of the study of population in parentheses.

| Clinical characteristic | Patient count |
| --- | --- |
| Total number of patients | 1,903 |
| Age<br>- 0-18 years<br>- 19-49 years<br>- 50-64 years<br>- ≥ 65 years | 50 (2.6%)<br>635 (33%)<br>522 (27%)<br>696 (36%) |
| Gender<br>- Male<br>- Female | 973 (51%)<br>930 (49%) |
| Race<br>- White<br>- Black<br>- Asian<br>- American Indian<br>- Other<br>- Unknown | 1,345 (71%)<br>217 (11%)<br>83 (4.4%)<br>67 (3.5%)<br>163 (8.6%)<br>41 (2.1%) |
| Ethnicity<br>- Non-Hispanic<br>- Hispanic<br>- Unknown | 1,546 (81%)<br>298 (16%)<br>71 (3.8%) |
| Comorbidities<br>- Anemia<br>- Asthma<br>- Cancer<br>- Chronic kidney disease<br>- Chronic obstructive pulmonary disease<br>- Heart failure<br>- Hyperglycemia<br>- Hypertension<br>- Obesity<br>- Obstructive sleep apnea<br>- Type 1 diabetes mellitus<br>- Type 2 diabetes mellitus | 556 (29.2%)<br>220 (11.6%)<br>402 (21.1%)<br>309 (16.2%)<br>147 (7.7%)<br>226 (11.9%)<br>263 (13.8%)<br>815 (42.8%)<br>479 (25.2%)<br>301 (15.8%)<br>84 (4.4%)<br>376 (19.8%) |

In order to capture the complications and comorbidities recorded in the clinical notes with positive sentiment, we ran the 1.1 million patient notes through a neural network based software that captures the sentiments of phenotypes from clinical notes.

For comorbidities, we considered 21 risk factors for COVID-19 severe illness reported by the CDC^4^, including: (1) anemia, (2) asthma, (3) BMI between 25-30 (overweight), (4) BMI between 30-40 (obese), (5) BMI >= 40 (severe obesity), (6) cancer, (7) cardiomyopathy, (8) chronic kidney disease (CKD), (9) chronic obstructive pulmonary disease (COPD), (10) coronary artery disease (CAD), (11) heart failure (HF), (12) hyperglycemia, (13) hypertension, (14) immunosuppressant medication usage, (15) liver disease, (16) neurologic conditions, (17) obstructive sleep apnea (OSA), (18) smoker (former or current), (19) steroid medication usage, (20) type 1 diabetes mellitus (T1D), (21) type 2 diabetes mellitus (T2D). We also note that bone marrow transplant, HIV/AIDS, pediatric conditions, pregnancy, sickle cell disease, solid organ transplant, and thalassemia were also considered, but were not included in the analysis because fewer than 20 patients had each of these comorbidities.

For complications, we considered 20 COVID-associated complications: (1) acute respiratory distress syndrome / acute lung injury (ARDS / ALI), (2) acute kidney injury (AKI), (3) anemia, (4) cardiac arrest, (5) cardiac arrhythmias, (6) chronic fatigue syndrome, (7) disseminated intravascular coagulation (DIC), (8) heart failure, (9) hyperglycemia, (10) hypertension, (11) myocardial infarction (MI), (12) pleural effusion, (13) pulmonary embolism (PE), (14) respiratory failure, (15) sepsis, (16) septic shock, (17) stroke / cerebrovascular incident, (18) venous thromboembolism / deep vein thrombosis (VTE / DVT), (19) delirium/encephelopathy, and (20) numbness.

A patient was determined to have a clinical phenotype (the comorbidity or complication, in question) if the clinical phenotype or synonyms were mentioned (with positive sentiment) within that patients’ electronic health records. For comorbidities, the mention must have occurred within a note at any point in the patient history prior to the patient’s first positive COVID-19 PCR test. Patients were only considered if they had at least one note within Mayo Clinic EHR system dated before days -31 relative to their first positive PCR test.

For the patients included in this study, we stratified the rates of new onset complications by comorbidities (**Table 2**). For each comorbidity, e.g. chronic kidney disease, we compare the rates of “new onset” complications in cohorts of COVID-19 patients with and without chronic kidney disease. To calculate the rate of a new-onset complication, e.g. acute kidney injury (AKI), the numerator is the number of patients with AKI recorded in the clinical notes (with positive sentiment) during but not prior to the time period. The denominator is the number of patients without AKI recorded in the clinical notes with positive sentiment prior to the time period.

**Table 2:** Overall rates of early onset complications stratified by comorbidity. In each row, we compare the rates of “early onset” complications in cohorts of COVID-19 patients with and without comorbidities, during the time period (Days 0-30) relative to the PCR diagnosis date. To calculate the rates of new onset complications, the numerator is the number of patients with any complication recorded in the clinical notes with positive sentiment during but not prior to the time period (see ***Methods*** for full list of complications). The denominator is the number of patients without the complication recorded in the clinical notes with positive sentiment prior to Day 0. The rows with statistically significant differences are highlighted in **green**. The columns are: **(1) Comorbidity:** Comorbidity that defines the cohorts, including chronic conditions which are risk factors for severe COVID-19 disease, **(2) Overall rate of new onset complications in cohort with the comorbidity:** Overall rate of new onset complications from Days 0-30 in the cohort of patients with the comorbidity. **(3) Overall rate of new onset complications in cohort without the comorbidity:** Overall rate of new onset complications from Days 0-30 in the cohort of patients without the comorbidity, **(4) Relative risk [95% C**.**I**.]: (Rate of complication in cohort with comorbidity) / (Rate of complication in cohort without comorbidity), along with the associated 95% confidence interval, **(5) BH-adjusted p-value:** Benjamin-Hochberg corrected p-value for the Fisher exact statistical significance test comparing the rates of overall complications in the cohorts of patients with and without the specified comorbidity.

| Comorbidity | Rate of new onset complication in cohort with the Comorbidity | Rate of new onset complication in cohort without the Comorbidity | Relative Risk [95% CI] | BH-adjusted p-value |
| --- | --- | --- | --- | --- |
| Hypertension | 148/351 (42%) | 60/1344 (4.5%) | 9.4 (7.1, 12) | 2.9E-64 |
| Chronic kidney disease | 68/167 (41%) | 140/1528 (9.2%) | 4.4 (3.5, 5.6) | 1.5E-22 |
| Type 2 diabetes mellitus | 73/204 (36%) | 135/1491 (9.1%) | 4 (3.1, 5) | 1.7E-20 |
| Cancer | 61/187 (33%) | 147/1508 (9.7%) | 3.3 (2.6, 4.3) | 1.5E-14 |
| Chronic obstructive pulmonary disease | 32/60 (53%) | 176/1635 (11%) | 5 (3.8, 6.5) | 1.6E-14 |
| Anemia | 61/195 (31%) | 147/1500 (9.8%) | 3.2 (2.5, 4.1) | 9.8E-14 |
| Coronary Artery Disease | 33/79 (42%) | 175/1616 (11%) | 3.9 (2.9, 5.2) | 3.1E-11 |
| Obstructive sleep apnea | 36/102 (35%) | 172/1593 (11%) | 3.3 (2.4, 4.4) | 9.6E-10 |
| Heart failure | 34/104 (33%) | 174/1591 (11%) | 3 (2.2, 4.1) | 2.9E-08 |
| Hyperglycemia | 27/85 (32%) | 181/1610 (11%) | 2.8 (2, 4) | 2.0E-06 |
| Immunosuppressant | 20/62 (32%) | 188/1633 (12%) | 2.8 (1.9, 4.1) | 4.1E-05 |
| Asthma | 22/88 (25%) | 186/1607 (12%) | 2.2 (1.5, 3.2) | 1.2E-03 |
| Cardiomyopathy | 13/43 (30%) | 195/1652 (12%) | 2.6 (1.6, 4.1) | 2.1E-03 |
| Smoker (Former or Current) | 31/152 (20%) | 177/1543 (11%) | 1.8 (1.3, 2.5) | 4.2E-03 |
| Type 1 diabetes mellitus | 6/16 (38%) | 202/1679 (12%) | 3.1 (1.7, 5.9) | 0.01 |
| Steroid | 60/377 (16%) | 148/1318 (11%) | 1.4 (1.1, 1.9) | 0.02 |
| Liver Disease | 5/16 (31%) | 203/1679 (12%) | 2.6 (1.3, 5.4) | 0.05 |
| BMI >= 40 (Severe Obesity) | 31/180 (17%) | 177/1515 (12%) | 1.5 (1.1, 2.1) | 0.05 |
| BMI >= 30, but <40 (Obesity) | 59/577 (10%) | 149/1118 (13%) | 0.77 (0.58, 1) | 0.08 |
| HIV | 1/1 (1e+02%) | 207/1694 (12%) | 8.2 (2.7, 14) | 0.13 |
| Stroke / Cerebrovascular incident | 4/21 (19%) | 204/1674 (12%) | 1.6 (0.73, 3.9) | 0.33 |
| BMI >= 25, but <30 (Overweight) | 55/453 (12%) | 153/1242 (12%) | 0.99 (0.74, 1.3) | 1 |

### Augmented curation to identify comorbidities and complications in clinical notes

An augmented curation approach was used to classify the sentiment of phenotypes mentioned in the clinical notes. In order to determine if a patient is indicative of a comorbidity or complication, we used a system consisting of:

- A high performance text retrieval and search engine that given a patient cohort consisting of one or more patients, pipes all the sentences, paragraphs and documents pertaining to those patients to a Named Entity Recognition (NER) system
- An NER system consisting of both a knowledge graph based entity recognition subsystem as well as a Bidirectional Encoder Representations from Transformers (BERT) based^5^ entity recognition system that, given any sentence or paragraph is able to identity whether it contains one or more phenotypes (comorbidities or complications etc.) so that all such sentences and paragraphs then are piped to a further downstream phenotype sentiment classification system.
- A BERT-based phenotype sentiment model^3^ that given a sentence or paragraph containing a phenotype/disease token provides accurately the sentiment as to whether or not the sentence indicates if the patient has the disease or not.
- A system that aggregates, patient-wise, all the sentence/paragraph sentiments, grouped by disease, and uses practical heuristics to determine if the patient may be confidently deemed to have the disease, based on the sentence level sentiments. As part of immediate future work, we are developing formal Bayesian models to aggregate the sentence level BERT sentiment information into a sound patient level sentiment inference.
- Presently we use heuristics based on the number of positive sentiment sentences found in a patient’s notes to deem whether or not the patient has a disease.

### Validation of the augmented curation model

In order to validate the augmented curation model for a set of phenotypes of complications/comorbidities, we manually labelled a set of 2,404 randomly selected sentences from the clinical notes containing the phenotypes. The true positive, true negative, false positive, and false negative rates are reported in **Supplementary Table S1**. Overall, the out-of-sample precision, recall, and accuracy values were 0.980, 0.982, and 0.966, respectively.

## Results

### Patient characteristics

1,903 patients were hospitalized with a diagnosis of COVID-19 between March 12, 2020 and October 15, 2020. Using the date of the first positive SARS-CoV-2 PCR test, we analyzed the clinical notes of each patient in their pre-COVID vs the post-COVID19 phase (**Figure 1A**). Using deep language models (**Figure 1B**) we extracted the 20 risk factors for COVID-19 severe illness reported by the CDC (**Figure 1C**) and the 18 COVID-associated complications (**Figure 1D**) in order to analyze their association in our cohort (**Figure 2-4**).

**Figure 1:**
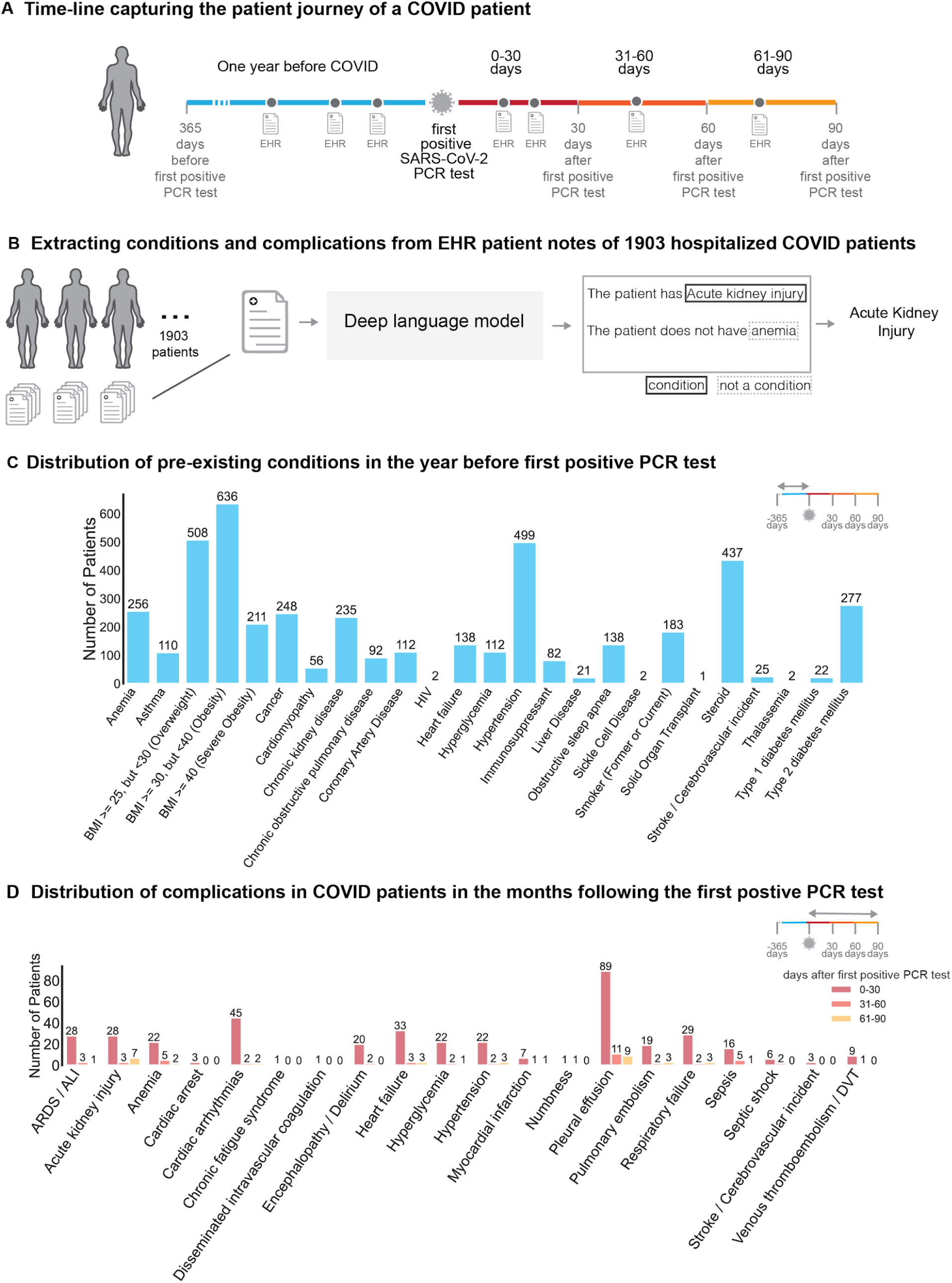
Study design and overview. **(A)** Relative timeline for each of the patients in the study, divided into the pre-COVID phase (1 year prior to the first positive PCR test), and the SARS-CoV-2 positive phase (90 days following the first positive PCR test). **(B)** Clinical notes from 1903 hospitalized patients are analyzed with a Bidirectional Encoder Representations from Transformers (BERT) model to extract the presence or absence of comorbidities and complications. **(C)** Distribution of pre-existing conditions in the first-year before the first positive PCR test **(D)** Distribution of complications at early (0-31 days), and late time points (31-60 days) (61-90 days).

**Figure 2.**
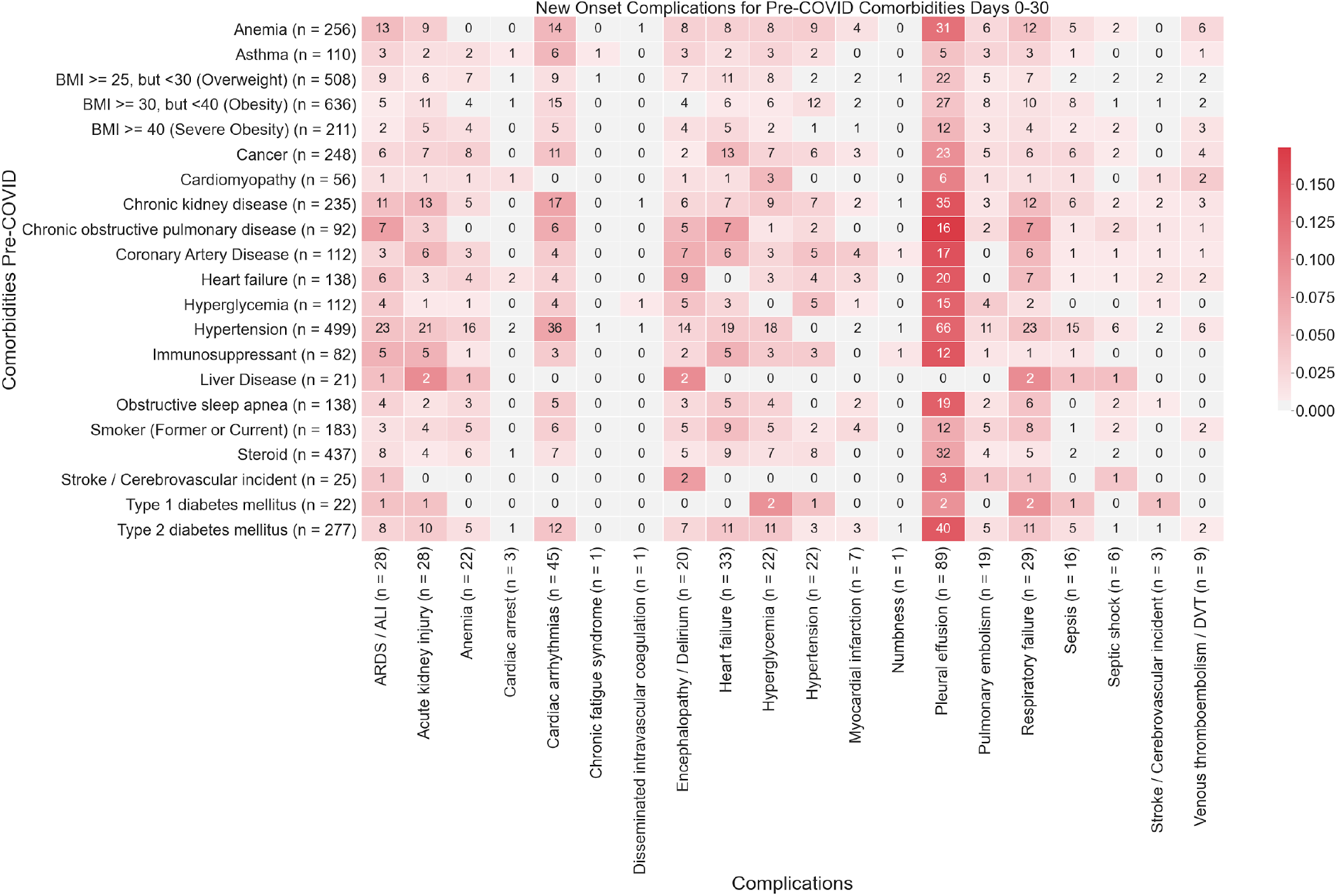
Heatmap showing associations between comorbidities and early-onset complications (0-30 days post-PCR) in COVID-19 patients. The color of each (comorbidity, complication) pair corresponds to (number of patients with the comorbidity that had the complication for the first time in the time window (0-30 days)) / (Total number of patients with the comorbidity). In particular, **darker shades of red** correspond to higher rates of complications, and **lighter shades of red** correspond to lower rates of complications. A patient is determined to have a comorbidity if it is recorded in their clinical notes with positive sentiment any time before their first positive PCR test. For each comorbidity row and for each complication column, the number of patients is shown in parentheses.

In **Table 1**, we present the general characteristics of the study population. All age groups are included, and, as expected from the severity of the disease in different age groups, more than 50% of the patients were over 50 year-old with only 2.6% of pediatric patients. Female, male and different ethnic origins of the US population are adequately represented. The most frequent comorbidities were hypertension (42.8%), diabetes/hyperglycemia (38%), obesity (25.2%) and cancer (21.1%), reflecting the most common causes of chronic diseases in the US.

The most common COVID complications recorded were respiratory (ARDS, respiratory failure, pulmonary embolism), followed by cardiovascular (hypertension, myocardial infarction, arrhythmia, stroke), acute kidney injury, anemia, sepsis and diabetic decompensation/hyperglycemia **(Figure 1D)**.

### Frequency of COVID-19 complications and association with underlying comorbidities

The main objective of our analysis was to identify the association between comorbidities and short-term (up to 30 days post-infection) and long-term (31-90 days post-infection) complications of COVID-19 infection. Here, we observe the majority of complications occur within the first month post-infection (**Figure 1D**).

We identify multiple comorbidities that are associated with significantly higher rates of any complications in the early onset time period (Days 0-30 post-PCR diagnosis). From this analysis, we validate that many of the CDC-reported risk factors for COVID-19 severe illness are associated with increased rates of early onset COVID complications across multiple organ systems (**Table 2**). Among these we identify hypertension (RR: 9.4, p-value: 2.9e-64) as the most significant risk factor followed by other cardiovascular chronic disease (heart failure, coronary artery disease, cardiomyopathy), anemia (RR: 3.2, p-value: 9.8e-14) and chronic kidney disease (RR: 4.4, p-value: 1.5e-22), as the most significant predictors of clinical complication in early COVID-19 infection.

### Respiratory complications

**Pleural effusions** are the most common early onset complications: 23% within the first months and 7% for ARDS/ALI, the second most frequent complication (**Figure 1D**). The primary risk factor for pleural effusion was hypertension (RR 9.2, p = 2.4e-22) **(Figure 2, Table 3)**. While the risk of new onset of pleural effusion is reduced after a month, our data reveal persistent risk of pleural effusion beyond 30 days post-infection, particularly among patients with type 1 diabetes (∼5%) **(Figures 3,4)**.

**Table 3:** Rates of early onset complications stratified by comorbidity. In each row, we compare the rates of “early onset” complications in cohorts of COVID-19 patients with and without comorbidities, during the time period (Days 0-30) relative to the PCR diagnosis date’. To calculate the rates of new onset complications, the numerator is the number of patients with the complication recorded in the clinical notes with positive sentiment during but not prior to the time period. The denominator is the number of patients without the complication recorded in the clinical notes with positive sentiment prior to the time period. Rows with cardiovascular complications are highlighted in **light red**, and rows with respiratory complications are highlighted in **light blue**. Rows are sorted first by complication, and second by statistical significance. The columns are: **(1) Complication:** Complication phenotype that is used to define the rates, including phenotypes associated with severe COVID-19 disease, **(2) Comorbidity:** Comorbidity that defines the cohorts, including chronic conditions which are risk factors for severe COVID-19 disease, **(3) Rate of new onset complication in cohort with the comorbidity:** Rate of complication from Days 0-30 in the cohort of patients with the comorbidity. **(4) Rate of new onset complication in cohort without the comorbidity:** Rate of complication from Days 0-30 in the cohort of patients without the comorbidity, **(5) Relative risk [95% C**.**I**.]: (Rate of complication in cohort with comorbidity) / (Rate of complication in cohort without comorbidity), along with the associated 95% confidence interval, **(6) BH-adjusted p-value:** Benjamin-Hochberg corrected p-value for the Fisher exact statistical significance test comparing the rates of the specified complications in the cohorts of patients with and without the specified comorbidity.

| Complication | Comorbidity | Rate of new onset complication in cohort with the Comorbidity | Rate of new onset complication in cohort without the Comorbidity | Relative Risk [95% CI] | BH-adjusted p-value |
| --- | --- | --- | --- | --- | --- |
| Acute respiratory distress syndrome / Acute lung injury | Hypertension | 23/489 (4.7%) | 5/1386 (0.36%) | 13 (4.8, 30) | 4.2E-08 |
| Acute respiratory distress syndrome / Acute lung injury | Anemia | 13/249 (5.2%) | 15/1626 (0.92%) | 5.7 (2.8, 12) | 2.9E-04 |
| Acute respiratory distress syndrome / Acute lung injury | Chronic kidney disease | 11/232 (4.7%) | 17/1643 (1%) | 4.6 (2.2, 9.6) | 2.7E-03 |
| Acute respiratory distress syndrome / Acute lung injury | Chronic obstructive pulmonary disease | 7/91 (7.7%) | 21/1784 (1.2%) | 6.5 (3, 15) | 2.7E-03 |
| Acute respiratory distress syndrome / Acute lung injury | Immunosuppressant | 5/77 (6.5%) | 23/1798 (1.3%) | 5.1 (2.2, 13) | 0.03 |
| Acute kidney injury | Hypertension | 21/412 (5.1%) | 7/1463 (0.48%) | 11 (4.5, 23) | 1.2E-07 |
| Acute kidney injury | Chronic kidney disease | 13/166 (7.8%) | 15/1709 (0.88%) | 8.9 (4.4, 18) | 4.1E-06 |
| Acute kidney injury | Type 2 diabetes mellitus | 10/229 (4.4%) | 18/1646 (1.1%) | 4 (1.9, 8.6) | 9.5E-03 |
| Acute kidney injury | Anemia | 9/190 (4.7%) | 19/1685 (1.1%) | 4.2 (2, 9.2) | 9.9E-03 |
| Acute kidney injury | Immunosuppressant | 5/59 (8.5%) | 23/1816 (1.3%) | 6.7 (2.9, 17) | 0.01 |
| Acute kidney injury | Coronary Artery Disease | 6/90 (6.7%) | 22/1785 (1.2%) | 5.4 (2.4, 13) | 0.01 |
| Anemia | Hypertension | 16/325 (4.9%) | 6/1556 (0.39%) | 13 (4.9, 30) | 4.3E-07 |
| Anemia | Cancer | 8/169 (4.7%) | 14/1712 (0.82%) | 5.8 (2.6, 14) | 3.7E-03 |
| Anemia | Heart failure | 4/68 (5.9%) | 18/1813 (0.99%) | 5.9 (2.3, 17) | 0.04 |
| Anemia | Chronic kidney disease | 5/114 (4.4%) | 17/1767 (0.96%) | 4.6 (1.9, 12) | 0.05 |
| Cardiac arrhythmias | Hypertension | 36/301 (12%) | 9/1557 (0.58%) | 21 (9.8, 40) | 2.7E-19 |
| Cardiac arrhythmias | Chronic kidney disease | 17/116 (15%) | 28/1742 (1.6%) | 9.1 (5.2, 16) | 2.3E-08 |
| Cardiac arrhythmias | Anemia | 14/134 (10%) | 31/1724 (1.8%) | 5.8 (3.2, 11) | 2.8E-05 |
| Cardiac arrhythmias | Chronic obstructive pulmonary disease | 6/47 (13%) | 39/1811 (2.2%) | 5.9 (2.9, 14) | 6.8E-03 |
| Cardiac arrhythmias | Type 2 diabetes mellitus | 12/180 (6.7%) | 33/1678 (2%) | 3.4 (1.8, 6.5) | 7.1E-03 |
| Cardiac arrhythmias | Cancer | 11/170 (6.5%) | 34/1688 (2%) | 3.2 (1.7, 6.3) | 0.01 |
| Cardiac arrhythmias | Asthma | 6/70 (8.6%) | 39/1788 (2.2%) | 3.9 (1.9, 9.2) | 0.04 |
| Encephalopathy / Delirium | Heart failure | 9/127 (7.1%) | 11/1756 (0.63%) | 11 (4.9, 26) | 3.9E-05 |
| Encephalopathy / Delirium | Hypertension | 14/461 (3%) | 6/1422 (0.42%) | 7.2 (2.7, 17) | 3.3E-04 |
| Encephalopathy / Delirium | Coronary Artery Disease | 7/104 (6.7%) | 13/1779 (0.73%) | 9.2 (3.9, 22) | 7.5E-04 |
| Encephalopathy / Delirium | Anemia | 8/231 (3.5%) | 12/1652 (0.73%) | 4.8 (2, 11) | 0.01 |
| Encephalopathy / Delirium | Chronic obstructive pulmonary disease | 5/85 (5.9%) | 15/1798 (0.83%) | 7.1 (2.9, 19) | 0.01 |
| Encephalopathy / Delirium | Hyperglycemia | 5/98 (5.1%) | 15/1785 (0.84%) | 6.1 (2.5, 17) | 0.02 |
| Heart failure | Hypertension | 19/384 (4.9%) | 14/1486 (0.94%) | 5.3 (2.7, 10) | 5.1E-05 |
| Heart failure | Cancer | 13/213 (6.1%) | 20/1657 (1.2%) | 5.1 (2.6, 10) | 4.2E-04 |
| Heart failure | Chronic obstructive pulmonary disease | 7/56 (12%) | 26/1814 (1.4%) | 8.7 (4.2, 19) | 5.0E-04 |
| Heart failure | Coronary Artery Disease | 6/55 (11%) | 27/1815 (1.5%) | 7.3 (3.4, 17) | 2.9E-03 |
| Heart failure | Type 2 diabetes mellitus | 11/206 (5.3%) | 22/1664 (1.3%) | 4 (2.1, 8.2) | 4.5E-03 |
| Heart failure | Smoker (Former or Current) | 9/157 (5.7%) | 24/1713 (1.4%) | 4.1 (2, 8.7) | 9.5E-03 |
| Heart failure | Immunosuppressant | 5/71 (7%) | 28/1799 (1.6%) | 4.5 (2, 12) | 0.04 |
| Hyperglycemia | Hypertension | 18/421 (4.3%) | 4/1460 (0.27%) | 16 (5.1, 40) | 1.9E-07 |
| Hyperglycemia | Type 2 diabetes mellitus | 11/187 (5.9%) | 11/1694 (0.65%) | 9.1 (4, 20) | 3.9E-05 |
| Hyperglycemia | Chronic kidney disease | 9/175 (5.1%) | 13/1706 (0.76%) | 6.7 (3, 15) | 9.3E-04 |
| Hyperglycemia | Anemia | 8/202 (4%) | 14/1679 (0.83%) | 4.7 (2.1, 11) | 0.01 |
| Hyperglycemia | Type 1 diabetes mellitus | 2/9 (22%) | 20/1872 (1.1%) | 21 (7.2, 73) | 0.03 |
| Hypertension | Chronic kidney disease | 7/33 (21%) | 15/1848 (0.81%) | 26 (12, 59) | 1.2E-06 |
| Hypertension | Anemia | 9/82 (11%) | 13/1799 (0.72%) | 15 (6.9, 34) | 3.3E-06 |
| Hypertension | Coronary Artery Disease | 5/27 (19%) | 17/1854 (0.92%) | 20 (8.6, 50) | 1.7E-04 |
| Hypertension | Hyperglycemia | 5/34 (15%) | 17/1847 (0.92%) | 16 (6.8, 41) | 4.5E-04 |
| Hypertension | Heart failure | 4/23 (17%) | 18/1858 (0.97%) | 18 (7.3, 49) | 1.3E-03 |
| Hypertension | Cancer | 6/121 (5%) | 16/1760 (0.91%) | 5.5 (2.3, 14) | 0.01 |
| Hypertension | BMI >= 30, but <40 (Obesity) | 12/458 (2.6%) | 10/1423 (0.7%) | 3.7 (1.6, 8.3) | 0.01 |
| Hypertension | Immunosuppressant | 3/34 (8.8%) | 19/1847 (1%) | 8.6 (3.2, 28) | 0.04 |
| Myocardial infarction | Coronary Artery Disease | 4/82 (4.9%) | 3/1814 (0.17%) | 29 (7.1, 1.1e+02) | 1.3E-03 |
| Myocardial infarction | Smoker (Former or Current) | 4/175 (2.3%) | 3/1721 (0.17%) | 13 (3.1, 50) | 0.01 |
| Myocardial infarction | Anemia | 4/235 (1.7%) | 3/1661 (0.18%) | 9.4 (2.3, 36) | 0.04 |
| Myocardial infarction | Heart failure | 3/117 (2.6%) | 4/1779 (0.22%) | 11 (2.9, 47) | 0.04 |
| Pleural effusion | Hypertension | 66/430 (15%) | 23/1384 (1.7%) | 9.2 (5.8, 14) | 2.4E-22 |
| Pleural effusion | Chronic kidney disease | 35/186 (19%) | 54/1628 (3.3%) | 5.7 (3.8, 8.4) | 9.8E-12 |
| Pleural effusion | Type 2 diabetes mellitus | 40/246 (16%) | 49/1568 (3.1%) | 5.2 (3.5, 7.7) | 9.8E-12 |
| Pleural effusion | Anemia | 31/200 (16%) | 58/1614 (3.6%) | 4.3 (2.9, 6.5) | 4.2E-08 |
| Pleural effusion | Heart failure | 20/98 (20%) | 69/1716 (4%) | 5.1 (3.3, 8) | 4.3E-07 |
| Pleural effusion | Chronic obstructive pulmonary disease | 16/73 (22%) | 73/1741 (4.2%) | 5.2 (3.3, 8.5) | 4.1E-06 |
| Pleural effusion | Coronary Artery Disease | 17/90 (19%) | 72/1724 (4.2%) | 4.5 (2.8, 7.4) | 1.5E-05 |
| Pleural effusion | Obstructive sleep apnea | 19/114 (17%) | 70/1700 (4.1%) | 4 (2.6, 6.5) | 2.2E-05 |
| Pleural effusion | Hyperglycemia | 15/87 (17%) | 74/1727 (4.3%) | 4 (2.5, 6.8) | 1.8E-04 |
| Pleural effusion | Immunosuppressant | 12/67 (18%) | 77/1747 (4.4%) | 4.1 (2.4, 7.2) | 7.9E-04 |
| Pleural effusion | Cancer | 23/211 (11%) | 66/1603 (4.1%) | 2.6 (1.7, 4.2) | 1.4E-03 |
| Pleural effusion | Steroid | 32/389 (8.2%) | 57/1425 (4%) | 2.1 (1.4, 3.1) | 0.01 |
| Pulmonary embolism | Hypertension | 11/478 (2.3%) | 8/1406 (0.57%) | 4 (1.6, 9.6) | 0.02 |
| Respiratory failure | Hypertension | 23/480 (4.8%) | 6/1394 (0.43%) | 11 (4.4, 25) | 8.5E-08 |
| Respiratory failure | Chronic kidney disease | 12/223 (5.4%) | 17/1651 (1%) | 5.2 (2.6, 11) | 6.9E-04 |
| Respiratory failure | Anemia | 12/241 (5%) | 17/1633 (1%) | 4.8 (2.4, 9.8) | 1.3E-03 |
| Respiratory failure | Chronic obstructive pulmonary disease | 7/83 (8.4%) | 22/1791 (1.2%) | 6.9 (3.2, 16) | 2.0E-03 |
| Respiratory failure | Type 2 diabetes mellitus | 11/266 (4.1%) | 18/1608 (1.1%) | 3.7 (1.8, 7.7) | 0.01 |
| Respiratory failure | Heart failure | 7/127 (5.5%) | 22/1747 (1.3%) | 4.4 (2, 10) | 0.02 |
| Respiratory failure | Smoker (Former or Current) | 8/179 (4.5%) | 21/1695 (1.2%) | 3.6 (1.7, 8.1) | 0.03 |
| Respiratory failure | Coronary Artery Disease | 6/108 (5.6%) | 23/1766 (1.3%) | 4.3 (1.9, 10) | 0.03 |
| Sepsis | Hypertension | 15/459 (3.3%) | 1/1428 (0.07%) | 47 (6, 1.7e+02) | 2.5E-07 |
| Sepsis | Chronic kidney disease | 6/206 (2.9%) | 10/1681 (0.59%) | 4.9 (1.9, 13) | 0.03 |
| Septic shock | Hypertension | 6/492 (1.2%) | 0/1405 (0%) | inf (2.1, 6.6e+02) | 2.9E-03 |
| Venous thromboembolism / deep vein thrombosis | Anemia | 6/233 (2.6%) | 3/1661 (0.18%) | 14 (3.6, 48) | 2.2E-03 |

**Figure 3.**
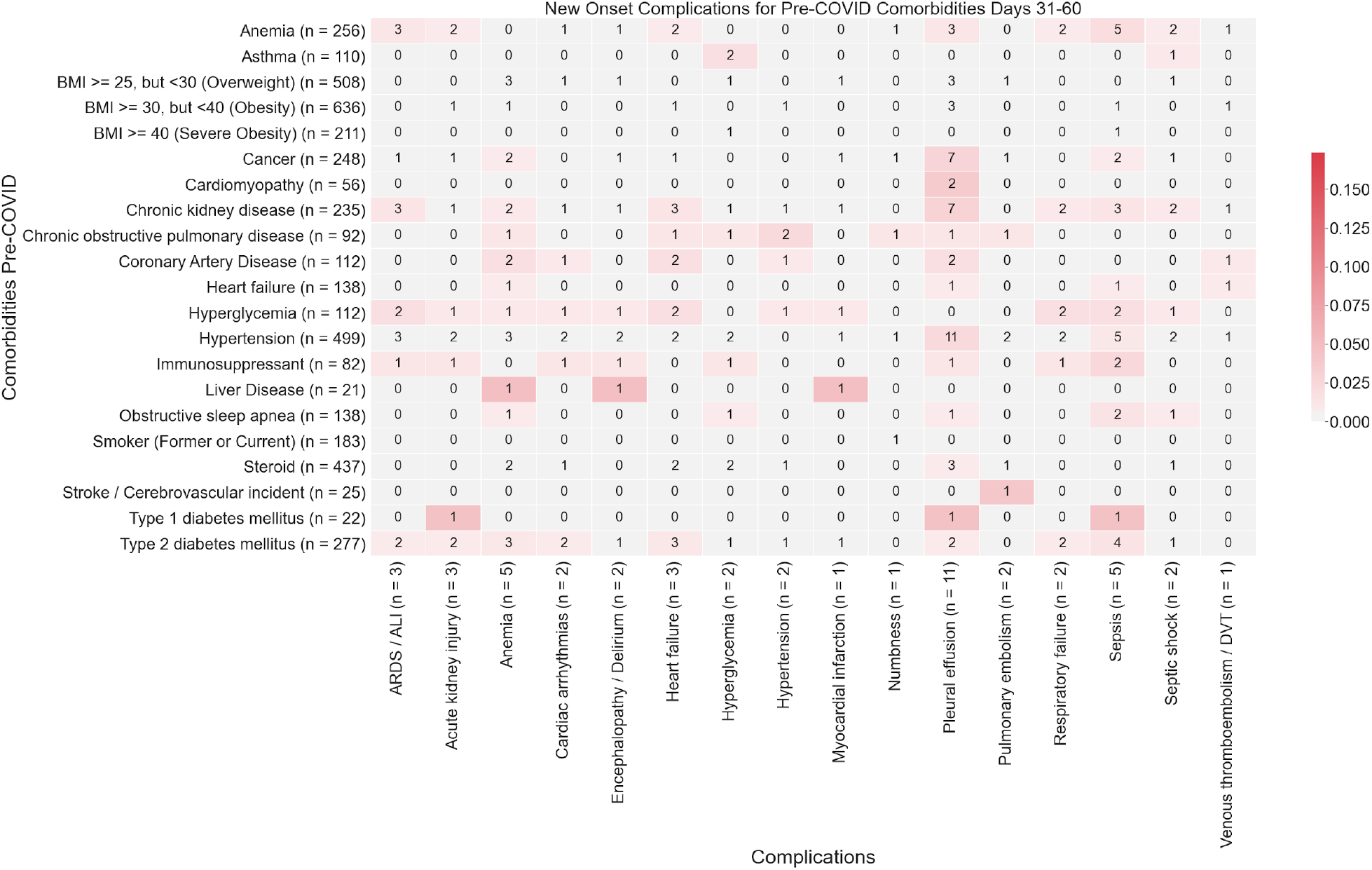
Heatmap showing associations between comorbidities and late-onset complications (31-60 days post-PCR) in COVID-19 patients. The color of each (comorbidity, complication) pair corresponds to (Number of patients with the comorbidity that had the complication for the first time in the time window (31-60 days)) / (Total number of patients with the comorbidity). In particular, **darker shades of red** correspond to higher rates of complications, and **lighter shades of red** correspond to lower rates of complications. A patient is determined to have a comorbidity if it is recorded in their clinical notes with positive sentiment any time before their first positive PCR test. For each comorbidity row and for each complication column, the number of patients is shown in parentheses.

**Figure 4.**
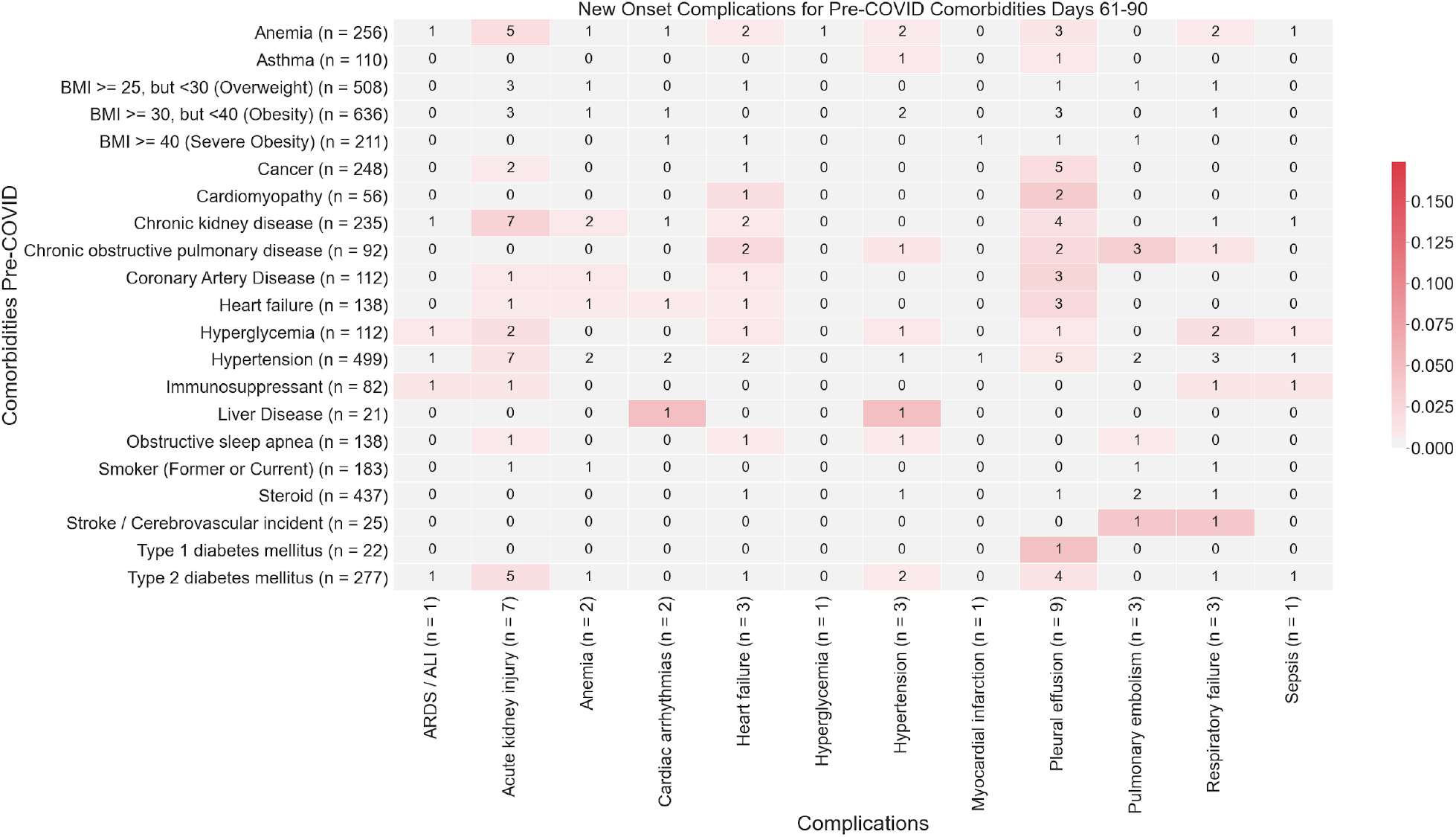
Heatmap showing associations between comorbidities and late-onset complications (61-90 days post-PCR) in COVID-19 patients. The number in each (comorbidity, complication) cell corresponds to the number of patients with the comorbidity that had the complication for the first time in the time window (61-90 days). The color of each (comorbidity, complication) pair corresponds to (Number of patients with the comorbidity that had the complication for the first time in the time window (61-90 days)) / (Total number of patients with the comorbidity). In particular, **darker shades of red** correspond to higher rates of complications, and **lighter shades of red** correspond to lower rates of complications. A patient is determined to have a comorbidity if it is recorded in their clinical notes with positive sentiment any time before their first positive PCR test. For each comorbidity row and for each complication column, the number of patients is shown in parentheses.

**Acute respiratory distress syndrome / acute lung injury** is the second most frequent and is the most dreaded complication of severe COVID-19 infection (7%) **(Figure 1D)**. In the early stages of COVID infection (i.e. 0-30 days post-infection), ARDS/ALI was most significantly associated with hypertension (p-value: 4.2e-8). The other most significantly associated baseline comorbidities include anemia (p-value: 2.9e-4) and chronic kidney disease (2.7e-3) **(Figure 2)**. In later stages of infection, we observe additional instances of ARDS/ALI, but at lower rates. Further, in later stages of infection, we fail to observe significant associations between baseline comorbidities and increased risk of new onset of ARDS/ALI (**Figures 3**,**4, Table 3**).

### Cardiovascular complications

**Cardiac arrhythmia** was the most common cardiovascular complication following COVID infection (12%) **(Figure 1D)**. Hypertension is by far the most important risk factor (RR= 21, p = 2.7e-19) **(Table 3)**. Up to 7% of hypertensive patients present with this complication within the first 30 days **(Figure 2)**. But the risk declines to less than 1% for new onset after one month post-infection **(Figures 3**,**4)**.

**Early onset COVID heart failure** is the second most common cardiovascular complication (9%) **(Figure 1D)**. It is primarily associated with coronary heart disease (RR=7.3, p=2.9e-3) and other cardiovascular risk factors (hypertension, anemia, type 2 diabetes, smoking) **(Table 3)**. But interestingly, cancer and immunosuppression are also uncovered as significant risk factors (RR=5.1 and 4.5, P = 4.0e-4 and 0.04, respectively). The cardiovascular complications examined in this study occur most frequently in days 0-30 post-infection **(Figure 2)**. Beyond 30 days, the risk of new-onset arrhythmia, hypertension, MI, PE/DVT dropped to less than 1% for all comorbidities **(Figures 3**,**4)**.

### Renal complications

**Acute kidney injury** is among the most common early-onset post-COVID complications (7%), (**Figure 1D**) and is associated in our cohort mostly with hypertension (RR=11, p=1.2e-7), and chronic kidney disease (RR 8.9, p=4.1e-6) **(Table 3) (Figure 2)**. Specifically, we observe acute kidney injury in 7% of hospitalized COVID patients in aggregate in early infection. The risk of acute kidney injury is highest in the early stages of infection (i.e. 0-30 days post-infection), while there is a reduction in the new onset of acute kidney injury beyond 30 days (either 31-60 days or 61-90 days) **(Figures 3**,**4)**.

### Neurologic Complications

**Encephalopathy and delirium** are a commonly observed complication of COVID (**c**), which is most associated in our cohort with heart failure (RR = 11, p = 3.9e-5), hypertension (RR = 7.2, p = 3.3e-4), and coronary artery disease (RR = 9.2, p = 7.5e-4) (**Table 3)**. Further, the risk of encephalopathy and delirium was observed to be highest in early COVID infection (**Figures 3**,**4**).

### Predictors of long-term complications of COVID-19 infection

While we observe a substantial reduction in the frequency of new onset of complications beyond 30 days post-infection, the risk of some complications is non-zero. In the case of pleural effusion, the risk decreases from 10-15% for all patients to less than 1% **(Figures 2**,**3**,**4)** with increased risk in patients with cardiomyopathy, chronic kidney disease, coronary artery disease, heart failure and hypertension. Patients with liver disease, stroke and type 1 diabetes also appear more susceptible to late-onset complications (days 31-90) **(Figure 3**,**4)**.

## Discussion

In the present study we have set out to understand the relationship between baseline comorbidities and clinical complications over the course of COVID-19 infection. Here, we leverage natural language processing of unstructured patient notes from 1,903 patients hospitalized with COVID-19 in the Mayo Clinic health system. While it stands to reason that individuals with poorer health status and multiple underlying comorbidities will experience worse outcomes during COVID-19 infection, our study reveals that not all risk factors are created equal and are associated with different complications. Previous studies have begun to uncover numerous factors associated with increased risk of more severe COVID-19 infection^6–10^ including hypertension, chronic kidney disease, type 2 diabetes, cardiovascular disease, and malignancy. In general, these studies have examined risk of severe COVID-19 infection, but have not examined the relationship between baseline comorbidities and risk of specific complications.

In our analysis, we observe that hypertension is the single most significant risk factor for all examined complications with exception of deep vein thrombosis. Notably, this is consistent with previous studies,, where patients with baseline hypertension have been reported to have higher risk of more severe COVID-19 disease^7,8^. Specifically, our data suggest that a recent history of hypertension is the strongest predictor of ARDS, the most significant and life-threatening complication of COVID-19, among hospitalized COVID-19 patients, similar to previous observations^10^. We further observed anemia, chronic kidney disease, immunosuppression, coronary artery disease and hyperglycemia to be associated with increased rates of ARDS. Our analysis further uncovered unexpected associations like between a history of cancer and immunosuppression with heart failure following COVID-19 infection.

Our data also highlight the temporal relationship between baseline health status and complications throughout COVID-19 infection. For example, cancer, obesity, and obstructive sleep apnea are associated with higher rates of short-term complications (days 0-30 post-PCR test), but not with late-onset complications. While many comorbidities are chronic (e.g. cancer, obesity, coronary artery disease and chronic kidney disease), others are amenable to short-term intervention, suggesting that tight control of modifiable risk factors might limit the risk of complication due to COVID-19 infection. For example, controlling hypertension, smoking cessation, treating anemia, and having tight glycemic control might reduce the rate of cardiovascular complications in the early stages of COVID-19.

Many of the comorbidities examined likely influence the development of complications, even in the absence of COVID-19 infection. For example, we do not observe new-onset pleural effusion among patients with pre-existing liver disease. It is possible that this is related to previous incidence of pleural effusion among patients with liver disease^11^. In the present analysis of the relationship between baseline co-morbidities and the likelihood of developing severe complications, we are not using a reference population of COVID-19 negative individuals. Instead, to understand these relationships, we have limited our inquiry to hospitalized COVID-19 patients and examined differences in the rates of clinical complications and various comorbidities. In future analyses, we will explore the rate of clinical complications in a control population of hospitalized COVID-negative patients to establish baseline complication rates within a hospitalized population.

At present, our analysis does not account for the co-dependent relationships between comorbidities or between complications. In many cases, individual patients likely have multiple complications, which can obscure the interpretation of data, particularly at later time points where we observe fewer events. Additionally, it is possible that many of the late-stage complications arise directly from baseline comorbidities rather than a direct result of COVID19 infection. This study can be leveraged for the development of controlled trials to identify appropriate prophylactic or therapeutic interventions for high-risk COVID-19 patients. Future analyses will focus on creation of a multivariate model to enable risk prediction of post-COVID complications^12^.

## Data Availability

Deidentified data will be made available upon reasonable request to corresponding author after peer-reviewed publication of this manuscript

## Supplementary Material

**Supplementary Table S1:**
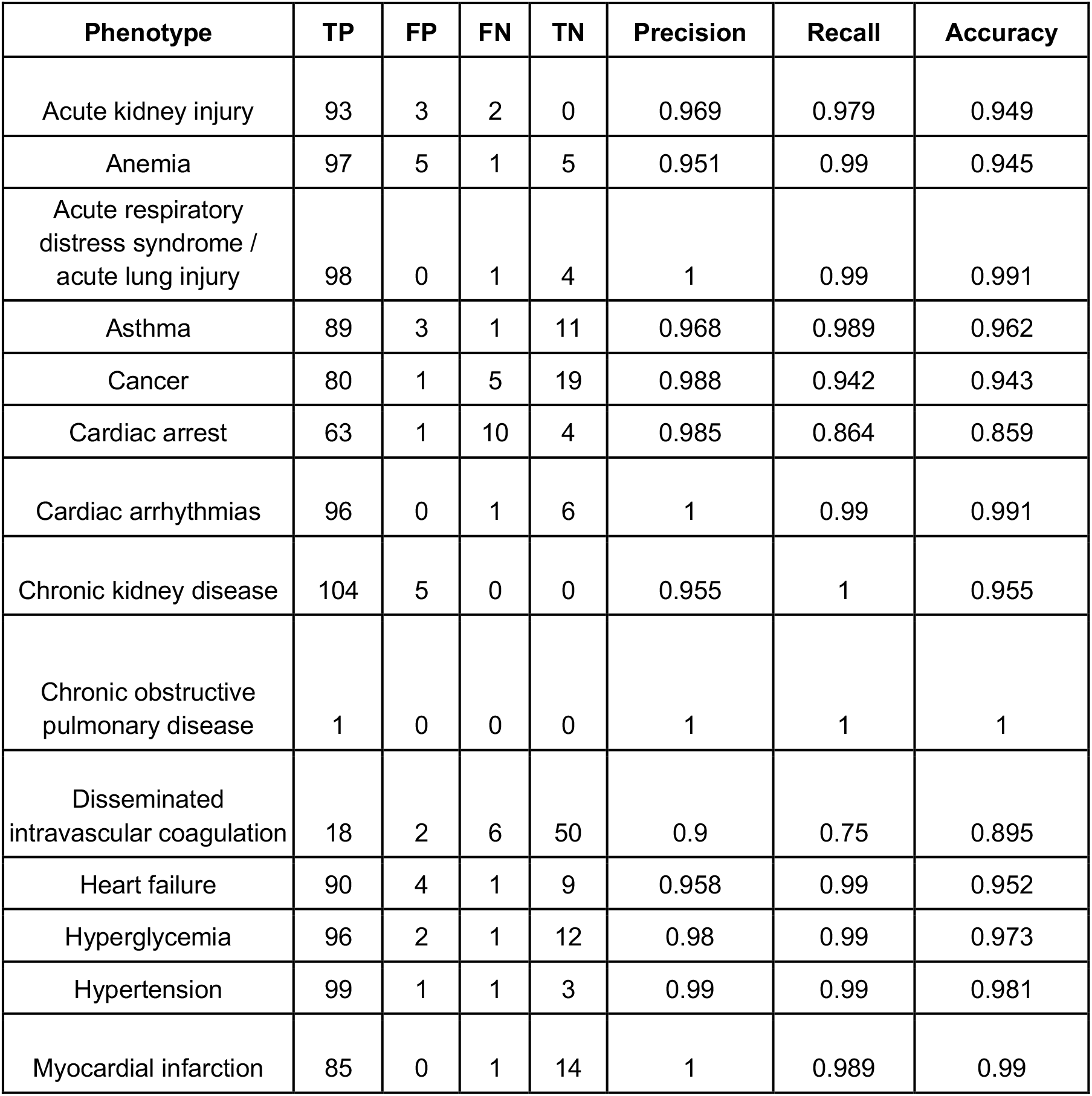
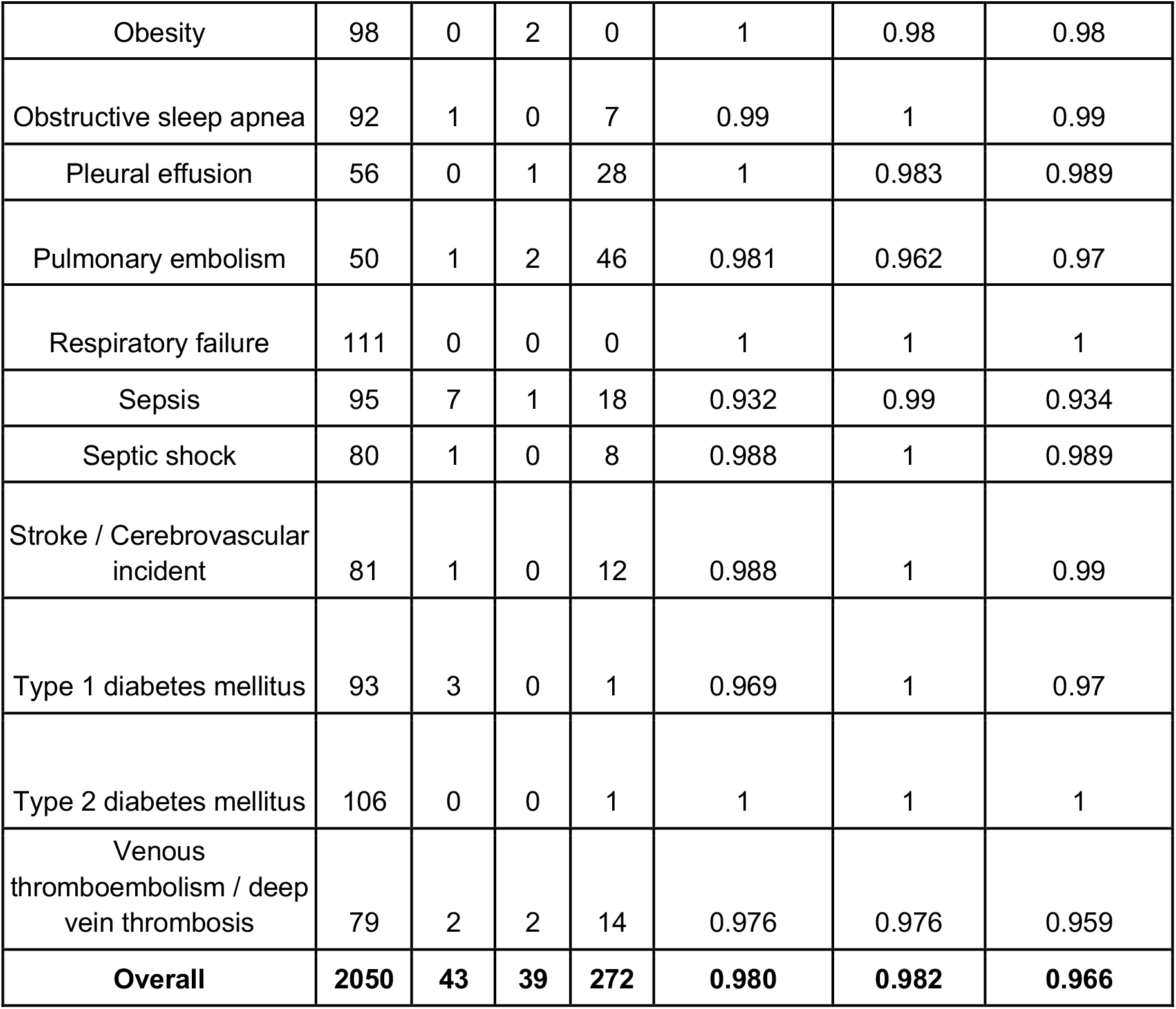
Sentence-level validation of BERT model to classify phenotype sentiment. The columns are **(1) Phenotype:** disease token identified in the sentence, **(2) TP (true positives):** count of sentences in which the BERT model correctly identified the sentiment as ‘Yes’, **(3) TN (true negatives):** count of sentences in which the BERT model correctly identified the sentiment as not ‘Yes’, **(4) FP (false positives):** count of sentences in which the BERT model incorrectly identified the sentiment as ‘Yes’, **(5) FN: (false negatives):** count of sentences in which the BERT model incorrectly identified the sentiment as not ‘Yes’, **(6) Precision:** precision of the BERT model, equal to TP/(TP+FP), **(7) Recall:** recall of the BERT model, equal to TP/(TP+FN), **(8) Accuracy:** accuracy of the BERT model, equal to (TP+TN)/(TP+TN+FP+FN).

| Phenotype | TP | FP | FN | TN | Precision | Recall | Accuracy |
| --- | --- | --- | --- | --- | --- | --- | --- |
| Acute kidney injury | 93 | 3 | 2 | 0 | 0.969 | 0.979 | 0.949 |
| Anemia | 97 | 5 | 1 | 5 | 0.951 | 0.99 | 0.945 |
| Acute respiratory distress syndrome / acute lung injury | 98 | 0 | 1 | 4 | 1 | 0.99 | 0.991 |
| Asthma | 89 | 3 | 1 | 11 | 0.968 | 0.989 | 0.962 |
| Cancer | 80 | 1 | 5 | 19 | 0.988 | 0.942 | 0.943 |
| Cardiac arrest | 63 | 1 | 10 | 4 | 0.985 | 0.864 | 0.859 |
| Cardiac arrhythmias | 96 | 0 | 1 | 6 | 1 | 0.99 | 0.991 |
| Chronic kidney disease | 104 | 5 | 0 | 0 | 0.955 | 1 | 0.955 |
| Chronic obstructive pulmonary disease | 1 | 0 | 0 | 0 | 1 | 1 | 1 |
| Disseminated intravascular coagulation | 18 | 2 | 6 | 50 | 0.9 | 0.75 | 0.895 |
| Heart failure | 90 | 4 | 1 | 9 | 0.958 | 0.99 | 0.952 |
| Hyperglycemia | 96 | 2 | 1 | 12 | 0.98 | 0.99 | 0.973 |
| Hypertension | 99 | 1 | 1 | 3 | 0.99 | 0.99 | 0.981 |
| Myocardial infarction | 85 | 0 | 1 | 14 | 1 | 0.989 | 0.99 |
| Obesity | 98 | 0 | 2 | 0 | 1 | 0.98 | 0.98 |
| Obstructive sleep apnea | 92 | 1 | 0 | 7 | 0.99 | 1 | 0.99 |
| Pleural effusion | 56 | 0 | 1 | 28 | 1 | 0.983 | 0.989 |
| Pulmonary embolism | 50 | 1 | 2 | 46 | 0.981 | 0.962 | 0.97 |
| Respiratory failure | 111 | 0 | 0 | 0 | 1 | 1 | 1 |
| Sepsis | 95 | 7 | 1 | 18 | 0.932 | 0.99 | 0.934 |
| Septic shock | 80 | 1 | 0 | 8 | 0.988 | 1 | 0.989 |
| Stroke / Cerebrovascular incident | 81 | 1 | 0 | 12 | 0.988 | 1 | 0.99 |
| Type 1 diabetes mellitus | 93 | 3 | 0 | 1 | 0.969 | 1 | 0.97 |
| Type 2 diabetes mellitus | 106 | 0 | 0 | 1 | 1 | 1 | 1 |
| Venous thromboembolism / deep vein thrombosis | 79 | 2 | 2 | 14 | 0.976 | 0.976 | 0.959 |
| **Overall** | **2050** | **43** | **39** | **272** | **0.980** | **0.982** | **0.966** |

